# Strengths and Limitations of Using ChatGPT: A Preliminary Examination of Generative AI in Medical Education

**DOI:** 10.1101/2025.03.12.25323842

**Authors:** Charlotte A. Taylor-Drigo, Anshul Kumar

## Abstract

**Introduction:** Objective Structured Clinical Examinations (OSCEs) are critical tools in medical education, designed to evaluate clinical competence by engaging students in tasks such as patient history-taking and physical examinations across multiple stations. Examiners assess student performance using structured rubrics to ensure objective evaluation. The COVID-19 pandemic spurred the adoption of technology in OSCEs, introducing virtual formats to maintain social distancing. While these advancements showcased the need for more interactive and realistic simulations, AI-driven tools like ChatGPT offer potential solutions. With its advanced conversational capabilities, ChatGPT can simulate patient interactions and provide real-time feedback, supporting active learning, experiential engagement, and cognitive management. Rooted in established learning theories, ChatGPT represents a promising avenue to enhance OSCEs by improving the evaluation of clinical competence and enriching training experiences.

**Method:** This pilot study engaged 20 faculty members who design and evaluate OSCE scenarios for medical students. Participants utilized ChatGPT in three simulated clinical cases that mirrored traditional OSCE formats. Following the simulations, feedback was collected via a survey to evaluate ChatGPT’s effectiveness and usability.

**Results:** The study found that participants appreciated AI as a standardized patient for its responsiveness, clarity, and ability to enhance clinical reasoning while reducing intimidation. However, key challenges included the lack of non-verbal communication, limited empathy, and an inability to perform physical examinations. Technical issues and inconsistent responses also affected the experience. While 20% of participants expressed interest in using AI in future OSCEs, they recommended a hybrid model combining AI with real standardized patients. This approach would leverage AI’s strengths while ensuring essential communication, empathy, and practical skills are effectively developed in clinical education.

**Conclusion:** Training ChatGPT to simulate diverse patient scenarios within OSCEs represents a significant innovation in medical education. By offering realistic simulations and precise feedback, ChatGPT has the potential to enhance both assessment accuracy and student preparedness for real-world clinical settings.

## Background

The Objective Structured Clinical Examination (OSCE) is a critical assessment tool in medical education, designed to evaluate the clinical competence of medical students. Developed by Harden and Gleeson (1979)1, the OSCE involves students rotating through structured stations where they demonstrate skills such as history-taking, physical examination, and patient communication. Examiners use checklists and rubrics to ensure a standardized and reliable evaluation of student performance.

The COVID-19 pandemic significantly disrupted traditional educational and healthcare models, accelerating the adoption of technology as an essential means of continuity. This shift underscored the role of digital innovations in medical education, including the adaptation of OSCEs to virtual platforms. According to Choi et al. (2020)4, virtual OSCEs emerged as a practical alternative to in-person assessments, enabling continued evaluation of students while adhering to social distancing guidelines. These virtual assessments, conducted via video conferencing, highlighted the need for more interactive and realistic simulations to maintain examination fidelity. Moreover, Twenge (2009)3 noted that Generation Me prefers structured yet interactive learning experiences, further emphasizing the necessity of technologically enhanced educational tools.

With technological advancements shaping medical education, artificial intelligence (AI) has gained prominence as a transformative tool in OSCEs. AI-driven applications (Khalil MK, Elkhider IA.2016)^9^, such as ChatGPT by OpenAI, offer innovative solutions for clinical education. These AI tools can simulate realistic patient interactions, provide real-time feedback, and support the development of clinical reasoning skills. The ability of ChatGPT to generate human-like responses makes it a suitable candidate for OSCE integration, enhancing both learning experiences and assessment accuracy.

Research supports the benefits of AI-based OSCE simulations. Maicher et al. (2019)^12^ demonstrated that virtual standardized patients effectively assess students’ information-gathering skills. Similarly, Ramchandani et al. (2024)^13^ reported positive student feedback on AI-driven OSCE simulations, emphasizing their role in improving clinical reasoning. AI-based simulations also address the learning preferences of millennial and Gen Z students, who favor flexible, technology-based resources (Hopkins et al., 2017)^2^. By offering on-demand, standardized patient interactions, AI enables students to practice at their own pace without scheduling constraints. Additionally, Wei Shen et al. (2024)^11^ found that AI-based simulations reduce student anxiety, fostering a safer environment for practicing clinical questioning and diagnosis.

Integrating ChatGPT into OSCEs aligns with several established educational theories. Constructivist Learning Theory, as discussed by Gandhi and Mukherji (2023)^7^, suggests that learners build knowledge through active engagement with content. ChatGPT facilitates this by allowing students to practice clinical scenarios interactively, with AI-driven scaffolding guiding their learning process. Experiential Learning Theory, developed by Kolb (1984)^10^, posits that knowledge emerges through hands-on experience. ChatGPT supports this by enabling students to engage in simulated clinical encounters, apply theoretical knowledge in practice, and reflect on their performance. Furthermore, Cognitive Load Theory (Sweller, 1988)^6^ asserts that instructional design should minimize extraneous cognitive load to optimize learning. ChatGPT contributes to this by breaking down complex cases into manageable segments and providing real-time clarifications, helping students focus on essential aspects of clinical reasoning without becoming overwhelmed.

This study aims to explore the training of ChatGPT to simulate lifelike patient interactions within OSCEs and assess its impact on medical education. It hypothesizes that integrating AI into OSCEs will enhance student learning experiences and improve the accuracy of clinical competence evaluations. By combining technological advancements with established educational theories, AI-driven OSCE simulations have the potential to revolutionize medical education, making assessments more accessible, flexible, and effective.

## Methods

This study employs a quantitative research design using a structured survey administered via “Qualtrics”. The study aims to measure specific variables related to the research question, allowing for statistical analysis to identify patterns, correlations, or differences among the study population.

### Ethics Approval

The Department Chair of the St. George’s University IRB approved the study and confirmed that the study conforms to all applicable guidelines and that all ethical matters were dealt with accordingly. A consent form was provided to all participating faculty in the study, which included information about the project. All consent forms were signed prior to the start of participation.

### Criteria for and Methods of Study Selection

#### Inclusion Criteria

Participants were adults aged 18 and above.

Participants were all proficient in English, as the survey was conducted in English. Participants had access to the internet to complete the online survey.

Participants were currently employed at St. George’s University (SGU) and participate in administering OSCEs to students in the PCM 501 course.

#### Methods of Study Selection

Participants were recruited based on availability and willingness. Invitations to participate were disseminated via the Clinical Instructors Year 2 course site on the Learning Management System (Sakai-used at SGU).

### Participants

The participants were faculty members currently employed at St. George’s University who assess students at OSCEs. A total of 20 participants were recruited. We chose a small sample size in the initial phase (pilot study) due to resource limitations, as we were using one account in ChatGPT to collect the data, which would be time-consuming with a larger cohort.

### OSCE Methodology

The OSCE is a comprehensive method used to assess the clinical competence of healthcare professionals. It involves a series of stations where candidates are tested on various skills. The OSCE aims to evaluate a wide range of clinical skills, including history-taking, physical examination, communication, clinical reasoning, and procedural skills. The history-taking and physical examination are conducted on a standardized patient (SP). A SP is an individual trained to consistently and accurately portray the characteristics of a real patient, including their medical history, symptoms, emotions, and personality. This role is pivotal in medical education and assessment because it allows healthcare students and professionals to practice and evaluate their clinical and communication skills in a controlled, realistic environment.

In this case, ChatGPT adopted the role of an SP. ChatGPT was trained using one account to function as a standardized patient using these steps (full documentation available in Appendix 1):

#### Data Collection of Medical Scenarios

The model was exposed to a variety of medical conditions, patient backgrounds, and scenarios for e.g., acute illnesses, chronic diseases, mental health issues, different emotions.

#### Preprocessing – Annotation

Responses were labeled indicating the type of scenario, the patient’s condition, and emotional tone.

### Model Training

#### Role-Play Training

The model was trained to understand the patient’s role by refining and using the prompt: “I would like you to behave as a standardized patient.”

#### Validation

The model’s responses were continuously validated to ensure that they align with current medical knowledge and best practices.

#### Scenarios Customization

(Scripts are outlined in Appendix 2): Participants used the customized scenarios in the same order each time:

Abdominal protocol

Musculoskeletal protocol

Psychiatry protocol

Each participant took about 60 minutes in one day to engage in the 3 cases and complete the survey. Each case consisted of a 17-minute session (15 minutes of encounter time and a 2-minute transition from one case to the other). Participants filled out a survey based on their experience doing an OSCE with ChatGPT.

### Materials and Procedures

In July 2024, 20 participants attended the Simulation Lab, located on the 3rd level of St. George’s Hall at St. George’s University. They performed the OSCE in the same sequence on a computer in room 21 of the lab. Each participant was allotted 15 minutes to interact with ChatGPT for each case, with a five-minute warning provided before the end of the session. Additionally, participants were given a two-minute transition between cases. This process was completed in one day. The survey was disseminated via email, and performance was ungraded.

### Measurement Methods and Data Collection Techniques

#### Survey Instrument

The survey was created using Qualtrics, a web-based platform that allows for the development of customized surveys with diverse question formats (e.g., multiple-choice, Likert scale, open-ended questions).

#### Data Collection Techniques

The survey was distributed via email. Participants completed the survey anonymously to promote honest and unbiased responses.

The survey remained accessible until the end of the study.

### Data Analysis Procedures

#### Data Cleaning

After data collection, responses were downloaded from Qualtrics in an Excel format.

Data was screened for incomplete responses or responses that did not meet the inclusion criteria, and those were removed.

#### Reporting

Results are presented in tables and pie charts for clear visualization of the data.

## Results

In this study, participants rated their experience with AI as a standardized patient in various ways (table 1): 35% rated it as excellent, 35% as good, and 30% as average. The most frequently mentioned benefits of AI as a standardized patient were its responsiveness and clarity, with participants appreciating the rapid and clear responses provided. The AI was also noted for its potential to help develop clinical skills, particularly in direct questioning and clinical reasoning. Additionally, participants felt more comfortable interacting with AI as a standardized patient compared to real patients, as it reduced feelings of intimidation.

**Table 1.** Quantitative feedback results.

| Question | Answer choices and results |  |  |  |  |
| --- | --- | --- | --- | --- | --- |
| How would you rate your overall experience with the AI standardized patient? | <u>Excellent</u><br>35% | <u>Good</u><br>35% | <u>Average</u><br>30% | <u>Poor</u><br>0% | <u>Terrible</u><br>0% |
| Would you prefer to have more AI standardized patient interactions in future OSCEs? | <u>Yes</u><br>20% | <u>Maybe</u><br>50% | <u>No</u><br>30% |  |  |
| In your opinion, which type of standardized patient contributes more to a student's learning experience in an OSCE setting? | <u>Both</u><br><u>equally</u><br>20% | <u>Real</u><br><u>patient</u><br>65% | <u>Artificial</u><br><u>intelligence</u><br>15% |  |  |

Regarding challenges, 85% of participants identified issues related to non-verbal communication, such as the inability to utilize non-verbal cues and limitations in performing physical examinations. Participants also highlighted difficulties in communication and rapport-building, citing a lack of empathy and proper clinical reasoning in AI as a standardized patient interaction. Technical and system issues were also noted, including system crashes and inconsistent responses. Participants expressed concerns about the realism and authenticity of AI as a standardized patient, stating that it was less realistic than interacting with a real patient, particularly in terms of communication and interpersonal skills.

Despite these challenges, 20% (table 1) of participants indicated interest in incorporating more AI in future OSCEs, acknowledging its potential to enhance clinical skills development, reduce pressure, and provide usefulness in specific scenarios that do not require physical examination. However, they noted that AI as a standardized patient should not be used solely for practical assessments but rather as part of a hybrid model alongside real standardized patients (table 1). This approach would help address the limitations of AI as a standardized patient particularly in developing communication skills and empathy.

Participants also emphasized the value of real standardized patients in contributing to a student’s learning experience. They highlighted the importance of real experiences, observation, and non-verbal communication in clinical reasoning and practical skills development. They also mentioned the limitations of AI, such as its scripted responses and inability to replicate non-verbal cues, while recognizing its potential role in patient management and scenario revision.

In conclusion, while AI as a standardized patient offers several benefits, especially in enhancing clinical reasoning and reducing pressure, its limitations, particularly in non-verbal communication and practical skills assessment, suggest that it is best used as part of a hybrid model alongside real standardized patients to provide a more comprehensive learning experience for students.

### Limitations of the study

Students were not included as participants in this study, as the focus was solely on faculty members involved in OSCE development and evaluation. This decision was made to gather expert insights on the use of ChatGPT in clinical simulations before involving students in subsequent stages. Additionally, transcripts of the simulated interactions were not analyzed in this study, though such an analysis could provide valuable insights into communication patterns and the quality of the simulations. Incorporating transcript analysis and student participation in future research could offer a more comprehensive understanding of ChatGPT’s effectiveness in enhancing OSCEs.

## Discussion

The introduction of practice sessions using ChatGPT for OSCEs enhances accessibility and provides a cost-effective opportunity for students to practice clinical encounters. This integration of technology into medical training aligns with the learning preferences of a new generation of students and offers several advantages:

### Standardized Assessment

ChatGPT ensures consistent and unbiased assessment of communication skills and clinical knowledge across students, minimizing examiner bias.

### Scalability

The use of ChatGPT allows for the simulation of patient interactions on a large scale, making it an ideal solution for institutions with a high number of medical learners.

### Flexibility

OSCE scenarios can be easily adapted and customized with ChatGPT to focus on specific learning objectives and competencies, providing tailored learning experiences.

### Immediate Feedback

Students benefit from immediate feedback on their performance, which supports continuous improvement in both communication skills and medical knowledge.

### Resource Efficiency

The implementation of ChatGPT reduces the reliance on standardized patients, which can be both costly and logistically challenging to manage.

### Data-Driven Insights

The data collected from student interactions with ChatGPT can offer valuable insights into areas where students may require additional training, allowing for targeted educational interventions.

Overall, the integration of ChatGPT into OSCEs presents a promising advancement in medical education, offering a scalable, flexible, and resource-efficient approach to enhancing student learning and assessment.

## Conclusion

The integration of AI language models, specifically training ChatGPT to act as a patient, represents a promising innovation in the realm of Objective Structured Clinical Examinations (OSCEs) within medical education. By incorporating ChatGPT, which can simulate a wide range of patient scenarios (enriched learning experience) and provide realistic responses, there is potential to enhance both the educational and assessment aspects of OSCEs (enhanced training effectiveness). This approach aims to improve the accuracy of skill assessments, offer diverse patient interactions, and ultimately better prepare medical students for real-world clinical settings.

## Supporting information

https://acrobat.adobe.com/id/urn:aaid:sc:VA6C2:c1563a23-b715-4a6e-b166-fa26ad623a14

https://acrobat.adobe.com/id/urn:aaid:sc:VA6C2:6d041978-d388-4ec2-b398-c126a8b3b4a4

https://acrobat.adobe.com/id/urn:aaid:sc:VA6C2:bea5b0f0-c2e5-4c2b-af67-484922654647

https://acrobat.adobe.com/id/urn:aaid:sc:VA6C2:1937e27a-fa51-43f6-b0db-082f722327ae

## Data Availability

All data produced in the present work are contained in the manuscript

## Notes

### Competing Interest Statement

The authors have declared no competing interest.

### Funding Statement

This study did not receive any funding

### Author Declarations

St. George's University IRB gave ethical approval for this work

