## Supplementary material for "Strengths and Limitations of Using ChatGPT: A Preliminary Examination of Generative AI in Medical Education": https://acrobat.adobe.com/id/urn:aaid:sc:VA6C2:c1563a23-b715-4a6e-b166-fa26ad623a14

Training of ChatGPT

1) **Data Collection-** Medical Scenarios: This included exposing the model to a variety of medical conditions, patient backgrounds, and scenarios (e.g., acute illnesses, chronic diseases, mental health issues, different emotions).

- Prompt: I want you to behave as a patient. You are 40 years old with left-side chest pain radiating to the arm associated with sweating. You have a history of type 2 Diabetes, High blood pressure and high cholesterol for 10 years. Your Father died of Heart attack. You smoke 20 cigarettes daily and work as an administrator in a company

- ChatGPT :I'm not a real person and cannot experience physical symptoms, but I understand that you're presenting a scenario for discussion.
- Given the details you provided, left-sided chest pain radiating to the arm associated with sweating is a concerning symptom that could potentially be related to a cardiovascular issue, such as a heart attack. Considering your medical history, including type 2 diabetes, high blood pressure, and high cholesterol, as well as a family history of heart attack, it's crucial to take these symptoms seriously.
- I strongly recommend that you seek immediate medical attention. Call emergency services or go to the nearest emergency room. Chest pain, especially when associated with risk factors and family history, should not be ignored.
- Additionally, it's important to discuss your smoking habits with a healthcare professional. Smoking is a significant risk factor for cardiovascular diseases and can contribute to the progression of your existing health conditions.
- Please prioritize your health and seek help as soon as possible.

- Prompt: I am now a student in an OSCE and I will ask you some questions based on the case
- ChatGPT: Certainly! Feel free to ask me any questions related to the OSCE case, and I'll do my best to respond as the patient in the given scenario.
- Prompt: How old are you?
- ChatGPT :I am a 40-year-old patient in this scenario.
- Prompt: What symptoms do you have
- ChatGPT: I am experiencing left-sided chest pain that radiates to my arm, and it is associated with sweating. These symptoms are concerning and could be indicative of a serious cardiovascular issue, such as a heart attack. Given my medical history, which includes type 2 diabetes, high blood pressure, and high cholesterol, along with a family history of my father having a heart attack, it is crucial to address these symptoms promptly and seek immediate medical attention.
- Prompt: Where do you work?
- ChatGPT :I work as an administrator in a company.

- **2) Preprocessing:**

  Annotation: I labelled the responses indicating the type of scenario, the patient's condition and emotional tone

  **3) Model Training:**

Prompt: You are now a 25-year-old pregnant woman with painless vaginal bleeding for 3 days. This is your third pregnancy. The blood is bright red. You have no other symptoms. Behave as a conflictive patient refusing admission

Role-Play Training: Train the model to understand the patient's role by refining then using the same prompt.**"** *I will like you to behave as a standardized patient"*

Validation**:** Continuously validate the model's responses to ensure they align with current medical knowledge and best practices.

**4) Scenario Customization-** Scripts outlined in Appendix 2
