## Supplementary material for "Strengths and Limitations of Using ChatGPT: A Preliminary Examination of Generative AI in Medical Education": https://acrobat.adobe.com/id/urn:aaid:sc:VA6C2:6d041978-d388-4ec2-b398-c126a8b3b4a4

**Scripts used:**

Script 1: Abdominal script

Presenting Complaint: “I’m having frequent diarrhea”

SP Name: Adrian Tuckman

CORE MEDICAL DETAILS

Student: Are there any other complaints you would like to discuss?

SP replies: There is no other complaint I would like to discuss at this moment.

SCRIPTED RESPONSES

Opening Statement, “I’m having frequent diarrhea”
Responses to Open-Ended Questions: For the past four days, I have been having frequent diarrhea I did not think much of it at first because I thought it was a simple stomachache from eating food on wall street, but my symptoms do not seem to be getting better.
Responses to Other Symptoms: I can also feel a “bump” on the back of the inner right cheek. Everything I touch with my tongue hurts. It also makes it difficult to chew my food. I am also having a crampy abdominal pain. I am losing weight because my clothes feel big. I also feel tired all the time.
Relieving Factors: The pain is sometimes relieved by defecation and 2 tablets of Tylenol
Aggravating Factors: None

Contingent Responses: If asked:

Can you describe your diarrhea? My stool is watery, and it is small amounts.
Do you see any blood in the stool? I have seen small amounts (about 1 teaspoon) of blood. The blood is not present in every stool.
How many episodes of diarrhea do you get in a day? I have 4-5 episodes in a day
Where is your abdominal pain? It is on the right side of my lower abdomen
Does your pain move / radiate anywhere? No, my pain stays in the same place, and it is there all the time
On a scale of 1 to 10, 10 being the worse, how would you rate this pain? The pain is a 7/10 intensity
How much weight did you lose? I went on the scale and noticed that I lost 20 pounds. My appetite has not changed, and I do not exercise.
Is there any change in your sleep pattern? I sleep throughout the night for 8 hours and still wake up feeling exhausted.

PAST MEDICAL HISTORY

Chronic Illness: I had similar experiences over the past year. The last time was 2 months ago. With previous episodes, diarrhea would last for 2 days and then stop.

Duration :1 year

Medications: Name: Tylenol
Dosage: 2 tabs 8 hourly
Duration: 4 days
Compliance: Good

Allergies: To medication -None
To environment -None

Preventative Health:

Vaccination: I had a set of vaccines at my last visit- I think influenza, tetanus, and pertussis. My doctor would know exactly which ones were given.
Date :1 year ago,
Last check Up: 1 year ago,
Findings: I was told I am in good health.
Sexual Health: Number of Partners -1spouse, Gender of Partners- Heterosexual, Protection- None

SIGNIFICANT FAMILY HISTORY

Paternal: My father has similar issues – his stool is bloodier with mucus. He has been seeing a gastroenterologist and is scheduled for a colonoscopy
Maternal: My mother has high blood pressure for which she takes medications
Siblings: None
Other relatives: None

SOCIAL HISTORY

Occupation: I am a salesclerk at Courts in Grand Anse

Education: Secondary school

Substance Abuse: Tobacco, type- Cigarettes Amount, 5 cigarettes per/day. Duration -10 years
Alcohol: Type, None
Illicit Substances: Type, None

Diet: I try to eat a balanced diet – it consists of chicken, fish, rice, provisions, and vegetables. Just before my diarrhea started, I ate food from a new vendor on wall street.

Exercise: Type, None

Physical examination: I have pain in the right lower quadrant

Impact of Disease:

Social - I have stopped going to work because my co-workers have noticed that I am using the restroom frequently and I feel very embarrassed. I must go to the bathroom so frequently; I cannot attend to the customers as I should.

Psychological: I am concerned that this is a serious issue like cancer. I always thought of growing old with my partner.

PERSONALITY PROFILE

Patient personality: Anxious and worried about the present situation

Reason for visit: I am worried about the diarrhea

Personal idea/s about illness: I think it may be the same disease as my father. My partner is also worried that this may be cancer because of the blood

Expectations for visit: I need relief of my symptoms so I can get back to work

Sources of concern: That this may be cancer

Questions: Can I take medication to stop the diarrhea? Is the blood in my stool a sign of cancer? Do you think that my symptoms can be cured?

Script 2: Musculoskeletal script

SP Name: Jane Park

CC: Opening Line: “I have pain in my right shoulder.

Student: Are there any other complaints you would like to discuss?

SP replies: There is no other complaint I would like to discuss at this moment.

Responses to Open-Ended Questions: I first noticed it two weeks ago whilst swimming but ignored it because I thought I might have pulled a muscle. It was not very painful at first but for the past week it’s been affecting my job, and I am not able to swim anymore.

Responses to Other Symptoms: My right shoulder has also been warm and swollen for one week.

Relieving Factors: Ibuprofen

Aggravating Factors: Moving my shoulder I also noticed the pain is worse at night especially when lying on my shoulder

Contingent Responses:

Can you describe the pain? Answer: The pain is dull and aching.

If you had to rate the pain on a scale of 0 to 10, 10 being the worst? Answer: The pain is a 5/10 intensity. If I move the arm, it becomes a 7/10. The pain is improved when I take ibuprofen, however the pain does not completely go away. With the medication the pain feels like a 2/10 intensity.

When was the last time you needed to take Ibuprofen? Answer: I took the ibuprofen just before I came to see you.

Does the pain move anywhere? Answer: The pain is always there (constant) and does not move anywhere. NOTE: If the student mentions rheumatoid arthritis, you will recognize this as the diagnosis of the arthritis you have in your hands.

PAST MEDICAL HISTORY

Chronic Illness: Name - Hypertension
Duration :1 year

Name: Arthritis of both my hands
Duration: 2 years

Medications:

Name: Methotrexate

Dosage :7.5mg once weekly
Duration: 2 years
Name: Lisinopril
Dosage :10 mg/day
Duration:1 year
Name: Water pill (diuretic)
Dosage :25mg/day
Duration :1 year
Name: Multivitamin
Dosage: 1/day
Duration: 2 years
Name: Ibuprofen
Dosage: 400mg
Duration: When pain
Compliance: Good

Allergies:

To medication- None
To environment-None
Preventative Health:
Vaccination -Unknown
Last check Up -3 months ago,
Sexual Health -Number of Partners, 1 for 10 years
Gender of Partners- Heterosexual
Protection- None
Type of Intercourse, Vaginal
STD testing:1 year ago
Result of testing: Negative,
OBGYN: Date of LMP, Menopause for 2 years
Sanitary Use: None
Menarche :12 years
Contraception: none
Pregnancies: 1
Obstetrical History: G1P1,
Hospitalization: Reason- Childbirth,
Duration - 2 days
Date - 24 years ago

SIGNIFICANT FAMILY HISTORY

Paternal: My father has hypertension and is controlled on medication,
Maternal: My mother is 70 years old has skin condition and arthritis. She is on medication.
Siblings: I have two younger brothers, and they are healthy,
Other blood-relatives: None

### SOCIAL HISTORY

Occupation: I am the manager of a hardware store
Education: University
Substance Abuse:
Alcohol – Type-Beers, Amount - 2 beers 3x per week for 10 years
Illicit Substances- Type, None
Diet: I try to maintain a good diet of vegetables, fish, and steam chicken but sometimes I indulge in salty foods.
Exercise :1. Walking Amount: ¼ mile daily 30 min 2/week, many years
2. Swimming,
Impact of Disease
Social: Inability to do work and care for grandson.
Psychological: The progressive nature of this pain over these couple of weeks is a source of worry and anxiety for me. Growing up I experienced first-hand the severity of pain my mother had to bear due to her longstanding arthritis. The thought of the possibility that I might have something similar would be hard to accept and deal with.

PHYSICAL EXAMINATION**:** Mild pain with all movements of the shoulder. Empty can test positive. All other special test negative

### PERSONALITY PROFILE

Patient personality: I am usually bold and outgoing
Reason for visit: I cannot do chores at home, and I have not been able to swim for about a week. I had to ask for time off work to come see you
Personal idea/s about illness: I think I may have injured it while swimming two months ago
Expectations for visit: I wish to be as efficient as I was before all this pain started
Sources of concern: I may not be able to swim again or have full function of my right shoulder
Questions: “Do you think my shoulder pain is related to my arthritis in my hands?” “If I take stronger pain medication, can I continue to swim?” “Do you think I damaged my shoulder while swimming?”

Script 3: Psychiatry script

SP Name: Camille Harry

SCRIPTED RESPONSES

Opening Statement: “I feel down”

Responses to Open-Ended Questions: I started feeling this way about 3 weeks ago. I don’t want to do anything or go anywhere. When I don’t need to be at work, I sleep half the day away locked in my room.

Responses to Other Symptoms: Its really difficult for me to get anything done. I don’t have enough energy, and I barely eat. I keep seeing things and I’m not sure if they are real.

Relieving Factors: None

Aggravating Factors: None

Contingent Responses:

How has your sleep been? I do not have enough energy to get things done even though I seem to be sleeping a lot more.

What do you mean when you say your concentration has changed? I am so down I can barely concentrate and get any work done.

Has your appetite changed? I have not had much of an appetite since this started.

Have you lost interest in certain activities that were once pleasurable? Lost interest in sex. Last intercourse 4 weeks ago

Have you had any hallucinations (seen things that weren’t real, or that no one else can see)? I think I’m losing my mind. I keep seeing my neighbor’s cat around the house, but they told me the cat died a few months ago. At first, I thought I was dreaming but I keep seeing it although I’m wide awake. I’m beginning to think the neighbors are lying and trying to trick me.

Have you had any thoughts of suicide (hurting yourself) or homicide (hurting others? No.
Do you often feel guilty or worthless? No.

PAST MEDICAL HISTORY

Chronic Illness: Name- Depression

Duration: For about five years, I feel down and sad from time to time. There isn't any specific trigger for my episodes of sad mood; it comes on, lasts a few weeks to a few months and then I go back to feeling normal. Each episode is very similar to the one I'm having right now. Five months ago, I had an episode where I was seeing and hearing strange things. I was eventually hospitalized and given treatment. At that time, I was not having any symptoms of depression. The entire episode lasted about three weeks.

Medications: Name- Since discharge I have not used any treatment. They gave me a prescription, but I did not fill it. I was supposed to return for a follow up appointment a few weeks after discharge, but I didn't. Unable to remember the name of medication.

Allergies:

To medication -None
To environment -None

Preventative Health:

Vaccination - Unknown
Last check Up: When hospitalized
Sexual Health -Number of Partners -1, Gender of Partners, Heterosexual, Protection , None , Type of Intercourse - Orovaginal ,
STD testing- Date: Never

SIGNIFICANT FAMILY HISTORY

Paternal: Alive and Well
Maternal: My mother has depression. She is on medication
Siblings: I have a sister who is healthy
Other relatives: I have an uncle who has schizophrenia

SOCIAL HISTORY

Occupation- Teacher

Education - Secondary

Substance Abuse - Tobacco, Type- None
Alcohol, Type- Wine, Amount- 5 glasses/week, 1/night Duration - 10 years
Illicit Substances-Type, None

Diet: I try to eat healthy; fresh fruits and vegetables that are in season. I like fish and chicken. I have not had much of an appetite since this started.

Exercise: type, None, I used to walk in the park but have not been feeling up to it recently,

Impact of Disease:

Social - I used to enjoy playing tennis and hanging out with my friends but for the last month I just wasn’t interested.

Psychological _I am very worried that if these symptoms don’t go away soon my marriage will end. I don’t even feel like having sex with him anymore. My friends have stopped asking me to go out anymore because every time they have asked in the last month, I said no. I am worried that I may lose my friends.

PERSONALITY PROFILE

Patient personality: You would describe yourself as dedicated to your family and friends.
Reason for visit: You have come to the office because you are worried their symptoms are getting worse. You hoped the symptoms would just go away, but now you are worried it could be something more serious
Personal idea/s about illness: I think it could be because I have not been taking my medication
Expectations for visit: I think I need a restart my meds so I am hoping you will write me another prescription
Sources of concern: I don’t want to have to take medication for the rest of my life. I was hoping to be able to stop taking meds eventually, but it seems like that will not happen

Questions: “Do you think that I’m crazy?” “Will I need to be hospitalized and medication?”
