## Supplementary material for "Strengths and Limitations of Using ChatGPT: A Preliminary Examination of Generative AI in Medical Education": https://acrobat.adobe.com/id/urn:aaid:sc:VA6C2:bea5b0f0-c2e5-4c2b-af67-484922654647

Table 1: What aspects of the AI standardized patient experience did you find most beneficial? (question 2)

Themes and examples excerpts

| Themes | Example excerpts |
|---|---|
| Development of Clinical Skills | Good for direct questioning. Can help to develop clinical reasoning especially information elicited during the history |
| | The information given was clear once the appropriate questions were asked |
| Comfort Level | Did not feel intimidated as I would with a patient |
| Responsiveness and Clarity | Rapid response |
| | Real time responses |
| | Quick responses but need to ask correctly to elicit an appropriate answer |
| | The responses were clear and concise |

Table 2: Would you prefer to have more AI standardized patient interactions in future OSCEs? (question 5)

Themes and examples excerpts

| Themes | Example excerpts |
|---|---|
| Benefits for Clinical Skills Development | Advancing clinical skills especially clinical reasoning |
| | Beneficial for extra training, revisiting scenarios, and cold cases in preparation for standardized patient encounter |
| | AI responses are generated quickly. I can ask another question while the response to the previous question is being generated |
| Limitations in Practical Assessments | Cannot be used solely for a practical assessment that involves performing a physical examination |
| | Nearly impossible to address communication skills and empathy using AI |
| | It will take a student longer to get the information if they must type their questions and read responses versus asking and actively listening |
| Hybrid System Suggestion | Maybe consider a hybrid system where they would use a simulated mannequin to perform the exam |
| | The option to have a percentage of AISPI in OSPE would help in comparison and perfecting the tool |
| Reduced Pressure and Enhanced Focus | There is less pressure as compared with a real patient |
| | Responses are generated quickly, allowing for follow-up questions |
| Usefulness in Specific Scenarios | Small cases not requiring physical examination |
| | Beneficial for extra training and revisiting scenarios |

Table 3: What challenges or limitations did you encounter while interacting with the AI standardized patient? (question 3)

| Themes | Example excerpts |
| --- | --- |
| Difficulty in understanding complex/ambiguous language | |
| Non-Verbal Communication Limitations | Unable to utilize non-verbal communication skills |
| | Limited in performing physical examination |
| Communication and Rapport | Lack of communication skill |
| | No rapport or bonding with AI |
| | Unable to show empathy or perform proper clinical reasoning |
| Technical and System Issues | System crashed and had incoherent responses |
| | AI did not respond to the prompts for the psychology case |
| Time and Interaction | Took more time to type than to ask questions |
| | Unable to do adequate closure with the patient |
| Inconsistent Responses | Management part: the AI responses varied sometimes acting as a doctor, sometimes as a patient, and sometimes provided information about the treatment/drugs instead of asking about how the drugs work or side effects |
| | Provided answers when prompted rather than allowing clinical reasoning |

Table 4: How did the AI standardized patient compare to a real standardized patient in terms of realism and authenticity? (question 4)

| Themes | Example excerpts |
|---|---|
| Limitations of AI | AI has limited memory in that it only knows what is put into it |
| | AI gives accurate information and reduces the need for clarifying information |
| | AI understands medical jargon while real patients struggle with it |
| Physical Examination | Physical findings and examination cannot be done |
| | Body language and physical is an important factor that may affect a patient's diagnosis which is limited in AI |
| Realism and Authenticity | Less realistic and authentic |
| | Some aspects of communication and interpersonal skills lack realism and authenticity such as showing empathy and using non-verbal communication skills |
| | Real encounters are much more interactive, and you are able to see the facial expression and body language of your patient |
| | Not the same as having a patient across from you whose body language in addition to responses can provide additional information |
| Interaction and Communication | Human responses in the initial encounter; distress, rapid speaking, broken sentences are not there |
| | "Responses quite similar to an actual standardized patient's response |
| | Not really comparing well. Responses are sometimes ambiguous |

Table 5: In your opinion, which type of standardized patient contributes more to a student's learning experience in an OSCE setting?

| Themes | Example excerpts |
|---|---|
| Value of Real Experiences | Real experiences are more valuable than simulated |
| | A real patient makes the clinical setting feel more real |
| | Human interaction involves more senses so memory formation is easier and more lasting than interacting with AI through a screen |
| Importance of Observation and Non-Verbal Communication | Part of the clinical reasoning process is to make observations of your patient from the time you walk in the room |
| | Observations during a conversation of a patient are important such as their speech, tone, painful distress, etc." |
| | Psychiatric patients are especially where observation and non-verbal communications are important |
| Practical Skills Development | Physical examination should be done and students must appreciate developing these practical skills rather than getting the findings immediately |
| | Verbalizing gives practice for the actual clinical world |
| Limitations of AI | There are limitations to AI with meeting the learning objectives |
| | AI makes the interview feel more like test-taking rather than a clinic |
| | The AI responses assist in calibrating the logarithms for best responses |
| Enhancement of Clinical Encounters | The personal experience in assessing non-verbal language in real patients enhances the encounter and may provide additional information |
