## Supplementary material for "Strengths and Limitations of Using ChatGPT: A Preliminary Examination of Generative AI in Medical Education": https://acrobat.adobe.com/id/urn:aaid:sc:VA6C2:1937e27a-fa51-43f6-b0db-082f722327ae

### Informed Consent

#### Welcome to the research study!

We are interested in understanding how a student feels doing an OSCE (Objective Structured Clinical Examination) using Artificial intelligence instead of a human standardized patient. For this study, your feedback is valuable in understanding the effectiveness of using Artificial Intelligence (AI) as a standardized patient compared to a real standardized patient. You will be asked to answer some questions about it. Your responses will be kept completely confidential.

The survey should take you around 10 minutes to complete. Your participation in this study is voluntary. You have the right to withdraw at any point during the study, without penalty. The Principal Investigator is Charlotte Taylor-Drigo who can be contacted at.

Thank you for your valuable input! Your feedback will assist us in continually improving the OSCE experience for all students in the PCM 501 Course.
By Clicking the button below, you acknowledge:

Your participation in the study is voluntary.
You are at least 18 years old.
You are aware that you may choose to terminate your participation at any time for any reason.

I agree

Program

School of Medicine

Other

What term are you facilitating?

Term 1

Term 2

Term 3/4

Term 5

How would you rate your overall experience with the AI standardized patient?

Terrible

Poor

Average

Good

Excellent

What aspects of the AI standardized patient experience did you find most beneficial?

What challenges or limitations did you encounter while interacting with the AI standardized patient?

Inability to detect sarcasm/humor.

Difficulty in understanding complex or ambiguous language.

Responses were inadequate.

Other

How did the AI standardized patient compare to a real standardized patient in terms of realism and authenticity?

Would you prefer to have more AI standardized patient interactions in future OSCEs?

No

Maybe

Yes

Please elaborate on your response.

In your opinion, which type of standardized patient contributes more to a student's learning experience in an OSCE setting?

Artificial Intelligence

Real Patient

Both equally

Please elaborate on your response.

### Block 1

Are there any additional comments or suggestions you would like to share regarding the use of AI or real standardized patients in OSCEs?

Powered by Qualtrics
